# Factors Associated with Malnutrition Among Under-Five Rohingya Refugee Children in Cox’s Bazar, Bangladesh: A Cross-Sectional Study

**DOI:** 10.1101/2025.09.22.25336301

**Authors:** Abdullah Safa Khan, Shimul Roy, Tamanna Tasmiah

## Abstract

**Objective:** To estimate the prevalence and determinants of malnutrition among under-five Rohingya refugee children living in Cox’s Bazar, Bangladesh.

**Design:** Community-based cross-sectional study.

**Setting:** Rohingya refugee camps, Cox’s Bazar, Bangladesh.

**Participants:** A total of 256 children aged 6–59 months residing in refugee camps were enrolled through convenience sampling.

**Primary outcome measures:** Stunting, wasting, underweight, and overweight, measured using WHO standards.

**Methods:** It was a cross-sectional study with a 256-sample size. Respondents were selected by convenience sampling. The target population was the Rohingya people of Cox’s Bazar, Bangladesh. A well-designed and pretested set of structured questionnaires was used to collect the information. The total study duration was 3 months of the period following the approval of the protocol. Informed consent was taken from the participants, and ethical issues were ensured by the Declaration of Helsinki. Data were analyzed by using SPSS.

**Results:** The prevalence of stunting, wasting, underweight, and overweight were 34.4%, 17.6%, 18.9%, and 6.9%, respectively. Age was inversely associated with stunting (AOR=0.97, 95% CI: 0.95–0.99, p=0.006). Male children had higher odds of wasting compared to females (AOR=2.03, 95% CI: 1.34–3.07, p=0.001). Children from poor households were more likely to be stunted (AOR=2.42, 95% CI: 1.31–4.48) and wasted (AOR=2.82, 95% CI: 1.23–6.47). Illiteracy among parents significantly increases the risks of malnutrition.

**Conclusions:** Malnutrition among Rohingya refugee children under five is alarmingly high. Both biological and socioeconomic determinants significantly influence nutritional status. Urgent multi-sectoral interventions addressing food security, parental education, and sanitation are required.

## Introduction

Child malnutrition remains one of the most pressing global health concerns, contributing to nearly half of all under-five deaths worldwide (WHO, 2021). Undernutrition manifests in stunting, wasting, and underweight, leading to impaired growth, increased susceptibility to infections, and long-term developmental deficits (Victora et al., 2010; Zlotkin et al., 2005). Refugee populations face heightened risks due to forced displacement, food insecurity, and poor healthcare access (Jayatissa et al., 2006; Wilkins et al., 2005). The Rohingya refugee crisis is among the largest in the world, with over 900,000 individuals residing in Cox’s Bazar, Bangladesh (UNHCR, 2022). Children make up more than half of this population, and malnutrition has been consistently reported as a major health burden (UNICEF USA, 2018). Despite several nutrition surveys, peer-reviewed studies focusing specifically on malnutrition determinants among Rohingya under-five children remain scarce. This study aims to fill this gap by estimating the prevalence and identifying associated factors of malnutrition.

## Methods

### Study design and setting

Cross-sectional study conducted in October–December 2023 in selected Rohingya camps, Cox’s Bazar, Bangladesh.

### Study population and sample

Children aged 6–59 months. A total of 256 were included, calculated using prevalence estimates (Rayhan & Khan, 2006; Jesmin et al., 2011).

### Data collection

Structured caregiver interviews collected data on demographics, parental education, occupation, food insecurity, and sanitation. Anthropometric measures were collected per WHO guidelines (WHO, 1995).

### Quality Control & Quality Assurance

The following steps were performed for quality control and assurance:

➢ Consistent assistance and direction from the supervisor.
➢ The researcher had collected and analyzed the data.
➢ An agreement was reached with the participants before gathering any data.
➢ A pretested questionnaire was utilized to gather data.

### Outcome measures

Stunting (HAZ < −2SD)

Wasting (WHZ < −2SD)

Underweight (WAZ < −2SD)

Overweight (WHZ > +2SD)

### Analysis

Descriptive statistics and multivariate logistic regression (SPSS v27).

### Ethics

Ethical approval obtained from North South University IRB. Written informed consent from caregivers.

## Results

The result of the study presents and explains the study’s conclusions as determined by data analysis. A complete sample of 256 respondents was appropriately evaluated and presented following the study’s objectives. The results are presented using figures and tables:

Table 1 shows the age distribution of the under-5 children, which shows that 40.6% from the 48– 59-month age group, 17.2% from 12-23 months, 14.5% from 24-35 months, 16% from 36-47 months, and 9.4% from 6-11 months.

**Table 1:**
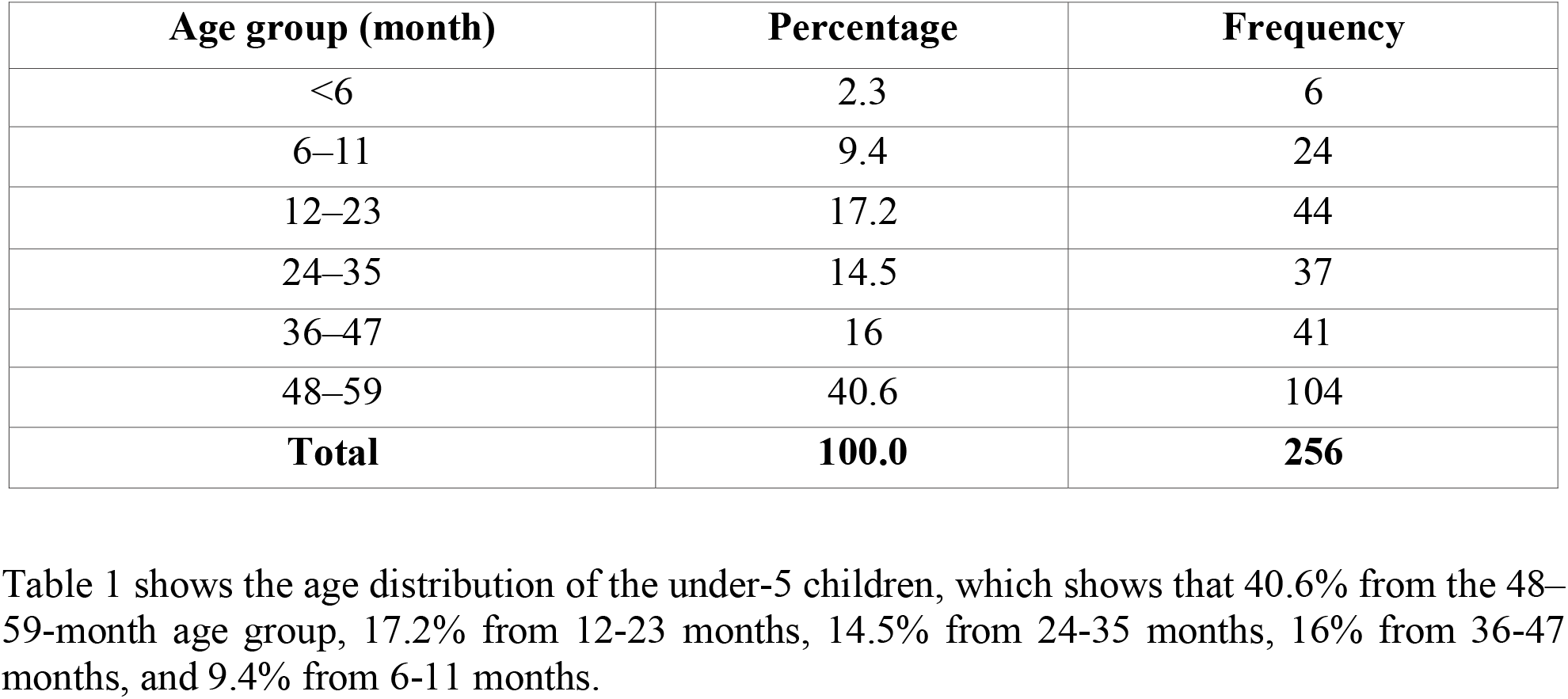
Age distribution of the respondents (n=256)

Table 2 displays the frequency of malnutrition and other sociodemographic details about the research participants. The children under five had a mean age of 37.22 months, and 56.6% of them were older than three years. Boys made up around 54% of the youngsters. The parents’ educational attainment was extremely low, with about 50% without a formal education. Nearly half of the dads (47.4%) worked throughout the day to provide for their families. It was discovered that fewer than half of the kids (45.5%) consumed at least four different kinds of meals. Poor bathroom facilities were identified in almost 36% of the homes. In the Rohingya refugee camps, stunting affected almost one-third (34.4%) of the children. 17.6% and 19% of the population, respectively, were underweight and wasting. Out of all the children in the camps, less than ten percent (6.9%) were overweight.

**Table 2:**
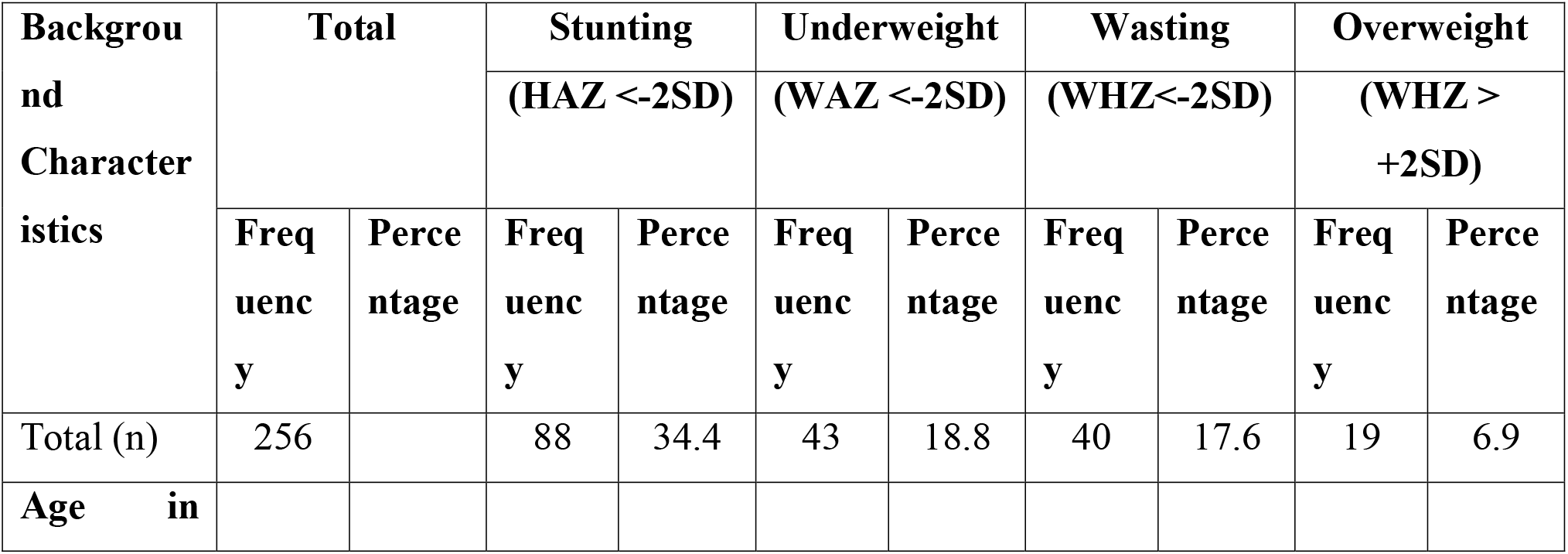
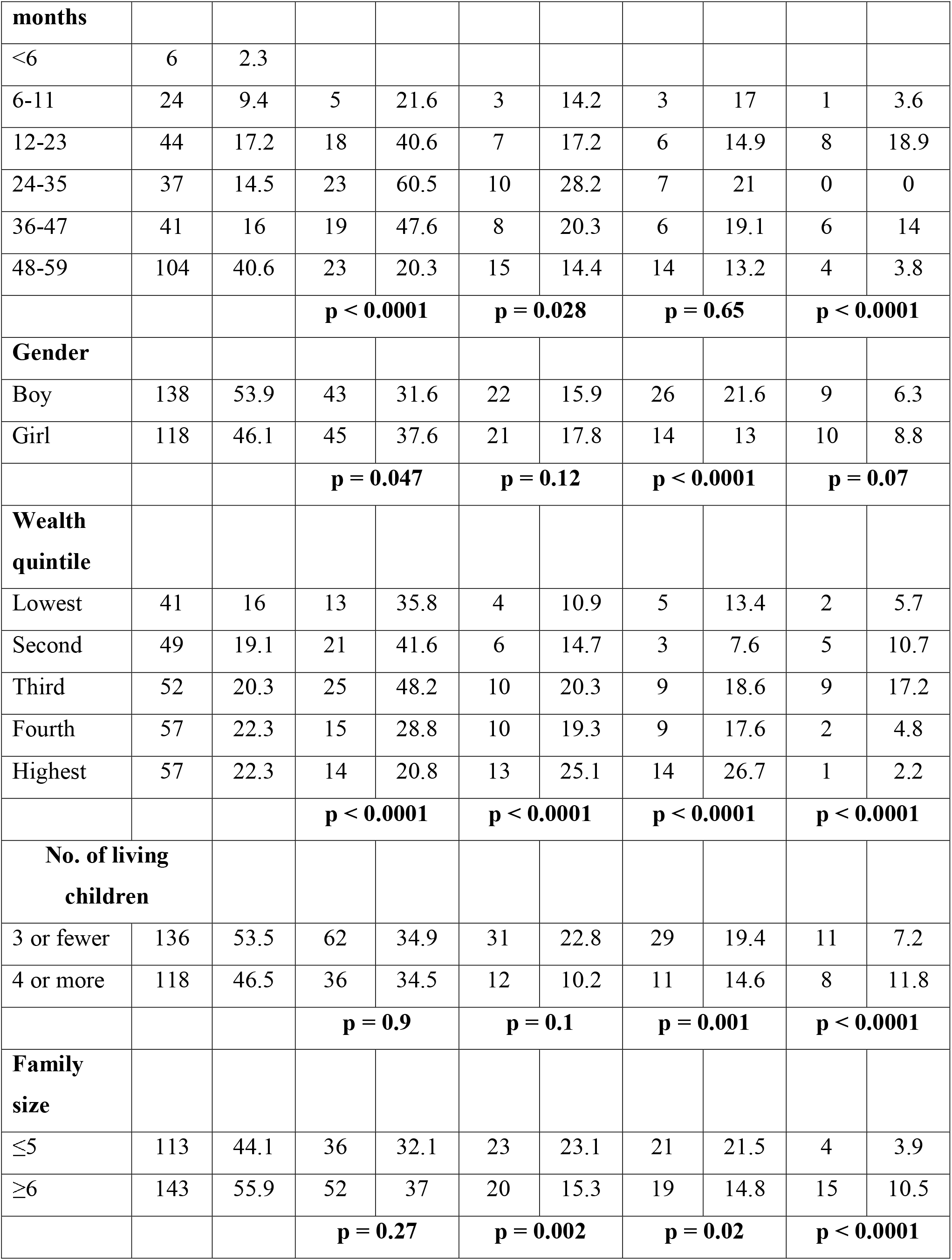
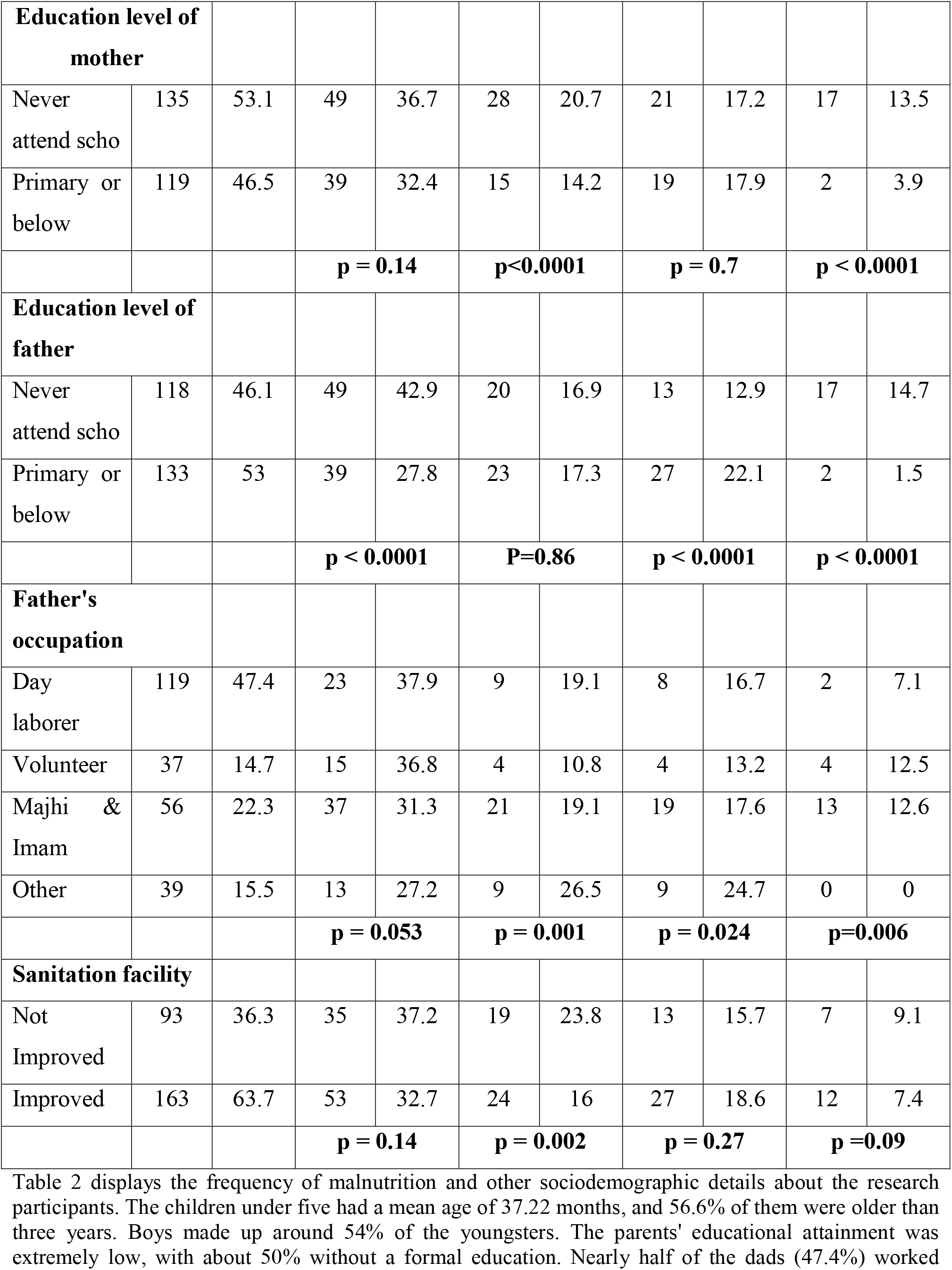
Association of Malnutrition with Socio-demographic Status of the Respondents (n=256)

The mean height of the male and female children was substantially greater (92.38 cm vs. 89.07 cm; p < 0.001), even though their mean weight (12.83 kg) was nearly equal (Table 5). The average HAZ, mean, and mean z-scores were, in that order, −1.34, −0.97, and −0.36. The mean WHZ was considerably larger in females (−0.20 vs. −0.56; p = 0.003), although the mean HAZ was lower in girls than in boys (−1.37 vs. −1.11; p = 0.01).

**Table 3.** Mean height, weight, age, and different nutritional status indicators of under-five children of Refugee Camps, Cox’s Bazar (n = 256).

| Indicator | Both Sex<br>Mean | Both<br>Sex SD | Boys<br>Mean | Boys<br>SD | Girls<br>Mean | Girls<br>SD | Mean<br>difference<br>(95% CI) | p-Value |
| --- | --- | --- | --- | --- | --- | --- | --- | --- |
| Age in month | 37.22 | 8.18 | 37.69 | 18.56 | 36.66 | 17.73 | 1.03 (-<br>1.13 3.18) | 0.35 |
| Height (cm) | 90.88 | 14.48 | 92.38 | 15.5 | 89.07 | 13.41 | 3.31 (1.56<br>5.06) | <0.001 |
| Weight (kg) | 12.83 | 3.51 | 12.98 | 3.8 | 12.16 | 3.29 | 0.82 (-<br>0.07 1.7) | 0.073 |
| MUAC (mm) | 151.1 | 10.89 | 151.6 | 10.37 | 150.1 | 11.45 | 1.49 (0.13<br>2.85) | 0.03 |
| Height for age<br>(HAZ) | -1.34 | 1.58 | -1.11 | 1.83 | -1.37 | 1.4 | 0.25 (0.05<br>0.45) | 0.01 |
| Weight for age<br>(WAZ) | -0.97 | 1.36 | -1 | 1.28 | -0.92 | 1.43 | -0.09 (-<br>0.26 0.07) | 0.26 |
| Weight for<br>height (WHZ) | -0.36 | 1.93 | -0.56 | 1.89 | -0.2 | 1.93 | -0.36 (-<br>0.59 -<br>0.13) | 0.003 |
| MUAC for age<br>(MUACZ) | -0.47 | 0.8 | -0.46 | 0.8 | -0.48 | 0.81 | 0.02 (-<br>0.08 0.11) | 0.76 |

*p-value from independent sample t-test.*

**Table 4:** Multiple logistic regression showing factors associated with stunting, wasting, and underweight.

| Variable | Stunting |  | Underweight |  | Wasting |  |
| --- | --- | --- | --- | --- | --- | --- |
|  | AOR<br>(95% CI) | p-value | AOR<br>(95% CI) | p-value | AOR<br>(95% CI) | p-value |
| **Age of child** | 0.97 (0.96<br>0.98) | 0.006 | 0.99 (0.98<br>1.00) | 0.2 | 1.01 (0.99<br>1.02) | 0.14 |
| **Gender** |  |  |  |  |  |  |
| Male | 0.82 (0.61<br>1.12) | 0.21 | 0.78 (0.52<br>1.15) | 0.21 | 2.03 (1.33<br>3.11) | 0.001 |
| Female (r) |  |  |  |  |  |  |
| **Wealth quintile** |  |  |  |  |  |  |
| Lowest | 1.08 (0.26<br>4.53) | 0.45 | 0.37 (0.15<br>0.87) | 0.024 | 2.88 (1.22<br>6.77) | 0.015 |
| Second | 1.63 (0.46<br>5.77) | 0.19 | 0.66 (0.3<br>1.44) | 0.29 | 1.15 (0.49<br>2.64) | 0.74 |
| Third | 2.42 (0.69<br>8.51) | 0.004 | 1.05 (0.53<br>2.07) | 0.87 | 3.0 (1.49<br>6.0) | 0.002 |
| Fourth | 0.78 (0.25<br>2.39) | 0.89 | 0.65 (0.34<br>1.28) | 0.21 | 1.51 (0.79<br>2.87) | 0.2 |
| Highest (r) |  |  |  |  |  |  |
| **Household monthly income** | 1.0 (1.0<br>1.0) | 0.28 | 1.0 (1.0<br>1.0) | 0.3 | 1.0 (1.0<br>1.0) | 0.003 |
| Birth order | 0.88 (0.66<br>1.17) | 0.49 | 1.00 (0.71<br>1.42) | 0.78 | 1.88 (1.28<br>2.77) | <0.0001 |
| No. of living children | 0.83 (0.57<br>1.22) | 0.32 | 1.81 (1.07<br>2.03) | 0.025 | 0.84 (0.49<br>1.45) | 0.44 |
| Family size | 1.17 (0.92<br>1.50) | 0.15 | 0.64 (0.46<br>0.90) | 0.02 | 0.81 (0.59<br>1.12) | 0.26 |
| **Education level of mother** |  |  |  |  |  |  |
| Never attend school | 0.74 (0.53<br>1.06) | 0.19 | 3.14 (1.11<br>8.53) | 0.001 |  |  |
| Primary or below (r) |  |  |  |  |  |  |
| **Education level of father** |  |  |  |  |  |  |
| Never attend school | 2.31 (1.74<br>3.68) | 0.03 |  |  | 0.41 (0.25<br>0.67) | <0.0001 |
| Primary or below (r) |  |  |  |  |  |  |
| **Father's occupation** |  |  |  |  |  |  |
| Day Laborer | 1.48 (0.87<br>2.54) | 0.34 | 0.31 (0.16<br>0.6) | <0.0001 | 0.32 (0.16<br>0.62) | 0.001 |
| Volunteer | 0.98 (0.54<br>1.79) | 0.91 | 0.16 (0.07<br>0.40) | <0.0001 | 0.41 (0.19<br>0.88) | 0.03 |
| Majhi & Imam | 1.22 (0.74<br>2.02) | 0.63 | 0.39 (0.21<br>0.74) | 0.001 | 0.52 (0.28<br>0.98) | 0.01 |
| Other (r) |  |  |  |  |  |  |
| **Toilet facility** |  |  |  |  |  |  |
| Not improved | 1.19 (0.54<br>2.6) | 0.67 | 1.56 (0.97<br>2.52) | 0.048 |  |  |
| Improved (r) |  |  |  |  |  |  |
| FIES score | 0.97 (0.87<br>1.08) | 0.69 | 0.96 (0.83<br>1.11) | 0.95 | 0.75 (0.64<br>0.88) | <0.0001 |
| Dietary Diversity Score of Children | 1.07 (0.87<br>1.33) | 0.56 | 1.03 (0.78<br>1.36) | 0.75 | 0.67 (0.49<br>0.84) | 0.007 |
| Mother's BMI | 1.01 (1.0<br>1.01) | 0.49 | 0.98 (0.97<br>1.0) | 0.055 | 0.98 (0.96<br>1.01) | 0.19 |
| FCS score | 1.01 (0.99<br>1.02) | 0.42 | 1.04 (1.02<br>1.06) | 0.001 | 1.02 (1.0<br>1.04) | 0.07 |

**Table 5.** Demographics of the study population.

| Variable | Value |
| --- | --- |
| Total children (n) | 256 |
| Mean age (months) | 37.2 ± 12.4 |
| Sex (Male) | 53.9% |
| Sex (Female) | 46.1% |
| Fathers with no formal education | ≈50% |
| Mothers with no formal education | ≈50% |
| Fathers working as day laborers | 47.4% |
| Households with poor sanitation | 36% |
| Households with food insecurity | 36% |

Table 6 presents the odds ratio (OR) and p-value for each factor’s contribution to stunting, wasting, and underweight in the multiple logistic regression model.

**Table 6.** Prevalence of malnutrition.

| Indicator | Prevalence (%) |
| --- | --- |
| Stunting | 34.4 |
| Wasting | 17.6 |
| Underweight | 18.9 |
| Overweight | 6.9 |

**Table 7.** Logistic regression results (key associations)

| **Factor** | **Associated outcome** | **AOR (95% CI)** | **p-value** |
| --- | --- | --- | --- |
| Child's age (months) | ↓ Stunting | 0.97 (0.95–0.99) | 0.006 |
| Male sex | ↑ Wasting | 2.03 (1.34–3.07) | 0.001 |
| Poor household (vs richest) | ↑ Stunting & wasting | 2.42–2.82 | <0.05 |
| Father illiterate | ↑ Stunting | 2.31 (1.08–4.92) | 0.03 |
| Mother illiterate | ↑ Underweight | 3.14 (1.58–6.26) | 0.001 |
| Poorest household (underweight) | ↓ Underweight | 0.37 (0.15–0.89) | 0.024 |

Children from middle-class quintile homes had 2.42 times greater risks of stunting than children from the wealthiest households (adjusted odds ratio, AOR = 2.42, p = 0.004). The odds of stunting were 2.31 times higher in children whose dads were illiterate than in children whose fathers had finished just elementary school (AOR = 2.31, p = 0.03). Nonetheless, the likelihood of stunting decreases as the child gets older (AOR = 0.97, p = 0.006).

Compared to girl children, boy children had 2.03 times greater odds of being wasted (AOR=2.03, p = 0.001). Compared to children from the richest homes, children from the poorest and middle-class households had 3.0- and 2.88 times greater risks of wasting, respectively. Compared to children whose dads had finished up to the elementary level of schooling, children whose fathers had never attended school had 59% reduced risks of wasting (AOR=0.41, p <0.0001). Compared to children whose father was employed in other jobs, children whose fathers worked in driving, business, or fishing had a 68%, 59%, and 48% lower chance of wasting. The FIES score (AOR=0.75, p <0.0001), the child dietary variety score (AOR =0.67, p =0.007), and the birth order (AOR=1.88, p <0.0001) were additional significant characteristics associated with wasting.

Children from the poorest homes had a 63% lower chance of being underweight than children from the richest households, in contrast to stunting and wasting (AOR = 0.37, p = 0.024). The OR for underweight rises as the number of alive children rises (AOR = 1.81, p = 0.025). In comparison to children whose mothers had finished up to the primary level of school, the odds of being underweight were 3.14 times greater in children whose mothers had no formal education (AOR = 3.14, p = 0.001). When FCS rises, so does the OR for underweight. A clean bathroom (AOR = 1.56, p = 0.048), a primary earner who goes fishing (AOR = 0.31, p < 0.0001), driving (AOR = 0.16, p < 0.0001), and business (AOR = 0.39, p = 0.001) were other significant sociodemographic characteristics associated with underweight.

### Demographics

Mean age = 37.2 months; 53.9% male. Half of the parents had no formal education. Food insecurity affected 36% of households.

### Malnutrition prevalence

Stunting (34.4%),

wasting (17.6%),

underweight (18.9%), and

overweight (6.9%).

### Determinants

#### Stunting

Associated with younger age (AOR=0.97, p=0.006), poor households (AOR=2.42, p=0.004), father’s illiteracy (AOR=2.31, p=0.03)

#### Wasting

More common in males (AOR=2.03, p=0.001) and poor households (AOR=2.82, p=0.015).

#### Underweight

Linked to maternal illiteracy (AOR=3.14, p=0.001).

## Discussion

This study confirms that malnutrition among Rohingya under-five children remains unacceptably high, consistent with prior reports from Bangladesh and other refugee contexts (Jesmin et al., 2011; Abudayya et al., 2007).

The inverse association between age and stunting underscores the importance of early-life nutrition (Victora et al., 2010). Higher wasting among boys aligns with previous findings of male vulnerability (Dasgupta et al., 2010). Socioeconomic determinants—parental education, poverty, and sanitation—were critical predictors (McKay et al., 2015; Methun et al., 2023).

Surprisingly, underweight was less common among the poorest, possibly due to targeted humanitarian aid (UNHCR, 2022).

Policy implications: Addressing malnutrition requires both nutrition-specific (breastfeeding, micronutrient support) and nutrition-sensitive (education, food security, WASH) interventions (Victora et al., 2016; Yousafzai et al., 2014).

## Limitations of This Study

The study’s sample size is inadequate, and the lack of coverage of a wider area may restrict the generalizability of its findings to the Rohingya community in the remaining camps. It was a cross-sectional study that examined only a single instance of nutritional status. Such information as the exact age of the children may be biased. The anthropometric measurements misrepresent the real health of the children. Self-reporting of a variable may introduce a degree of informational bias. Nonetheless, a reliable sampling procedure and sample size that is statistically justifiable could confirm the study’s conclusions.

## Conclusion

Malnutrition prevalence among Rohingya refugee children under five is alarmingly high, with clear links to biological and socioeconomic factors. Urgent, multi-sectoral interventions are needed to reduce the burden.

## Recommendations

✓ Strengthen maternal education and awareness on nutrition.
✓ Scale-up breastfeeding and complementary feeding promotion.
✓ Enhance food security programs with equity-focused targeting.
✓ Improve water, sanitation, and hygiene (WASH) facilities in camps.
✓ Implement regular monitoring and nutrition surveillance.

## Data Availability

All data produced in the present work are contained in the manuscript.

## Abbreviation

SAM: Severe Acute Malnutrition
MAM: Moderate Acute Malnutrition
MUAC: Mid-Upper Arm Circumference
FDMN: Forcefully Displaced Myanmar National
BMI: Body Mass Index
OTP: Outpatient Therapeutic Center
ISCG: Inter Sector Coordination Group
WHO: World Health Organization
SDG: Sustainable Developmental Goal
ORS: Oral Rehydration Salt
UN: United Nations
GAM: Global Acute Malnutrition
HAZ: Height-For-Age Z-Score.
SD: Standard Deviation.
WAZ: Weight-For-Age Z Score.
WHZ: Weight-For-Height Z Score.
AOR: Adjusted Odds Ratio.
CI: Confidence Interval.
FCS: Food Consumption Score;
FIES: Food Insecurity Experience Scale.

## Author contributions

The corresponding author wrote and reviewed the entire manuscript.

## Data sharing statement

Data will be available on demand.

---

**Figure 1.**
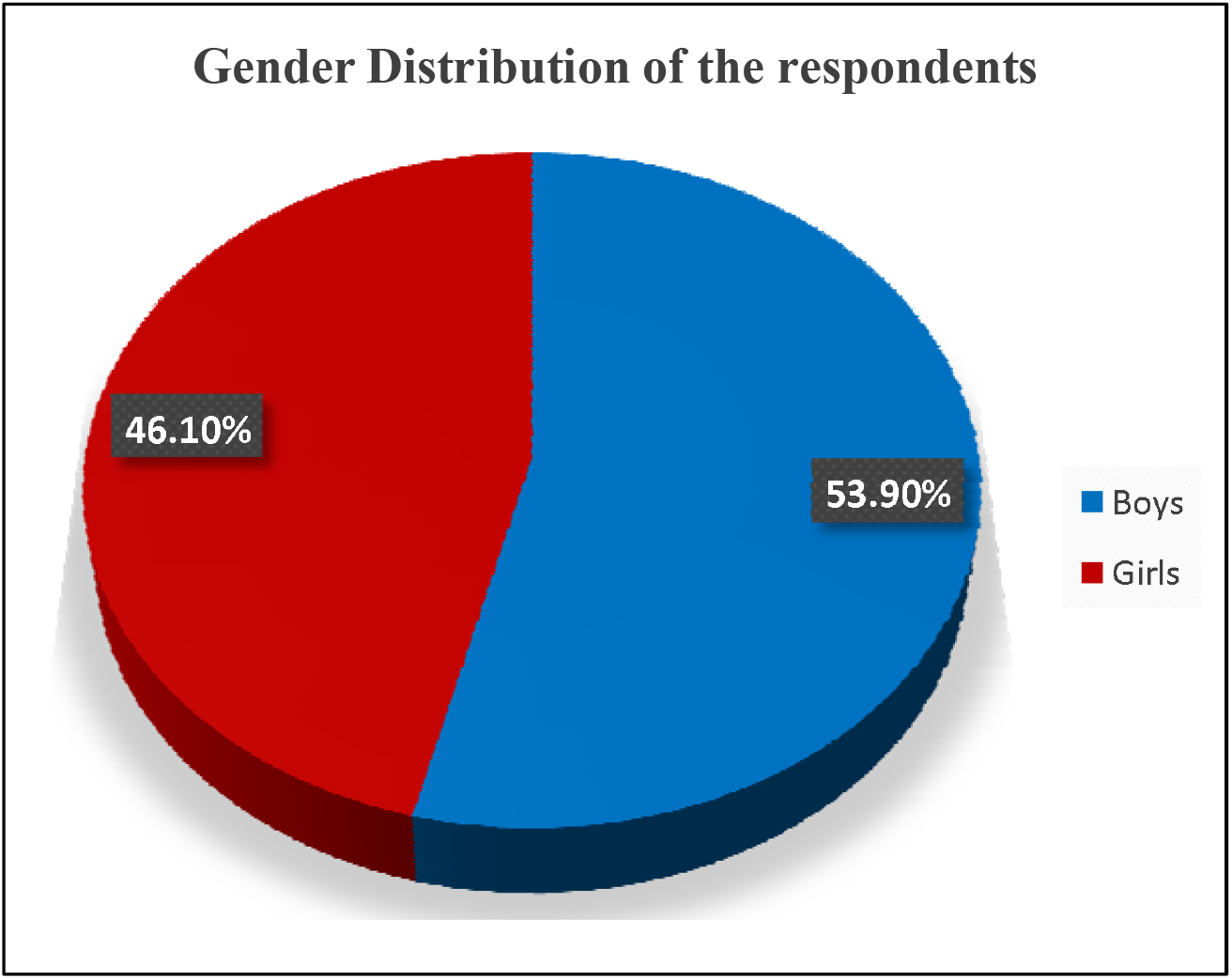
Gender Distribution of the respondents (n=256). The gender distribution of the children, boys and girls, was 53.90% and 46.10%.

**Figure 2.**
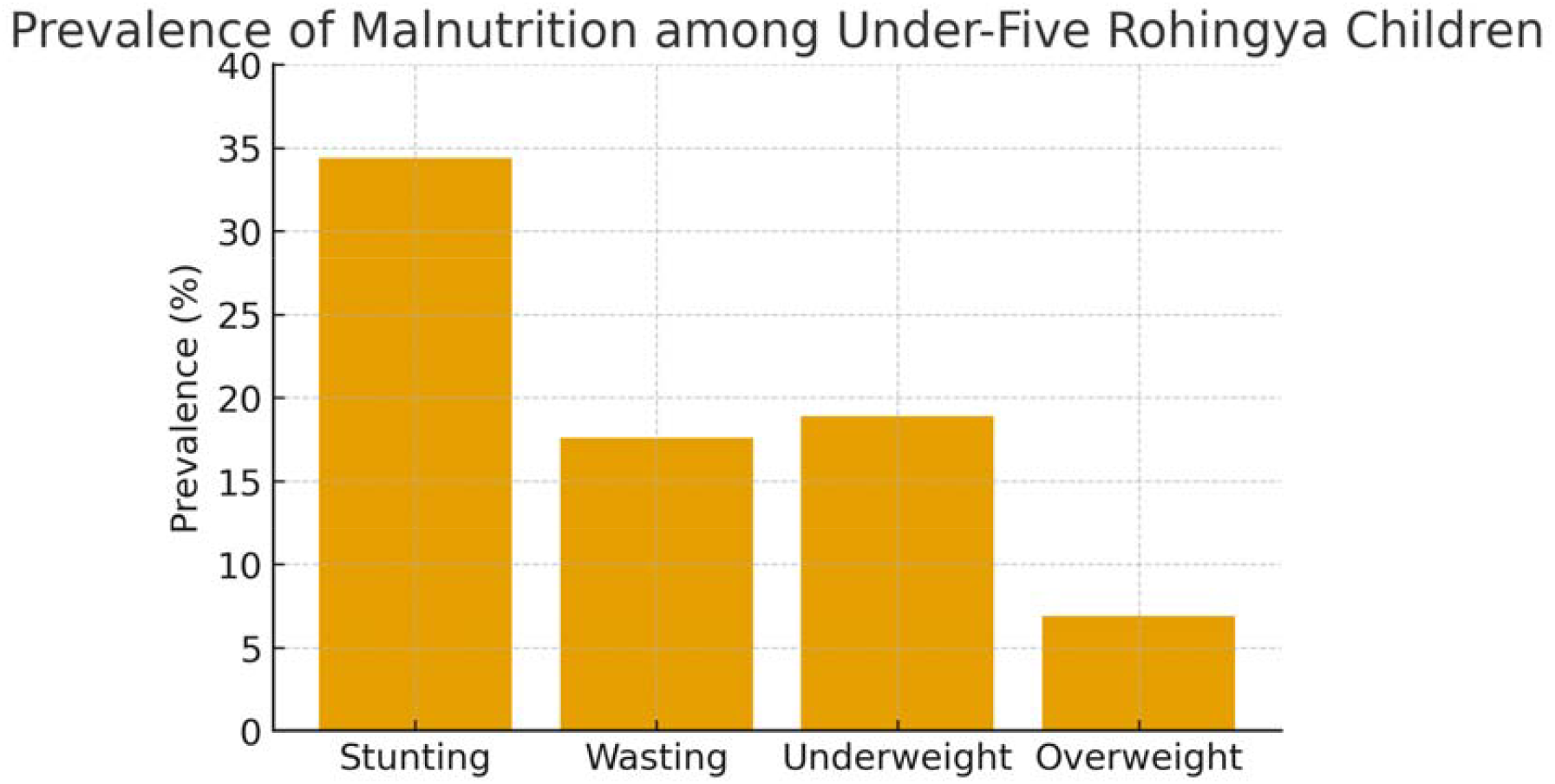
Prevalence of malnutrition among under-five Rohingya children

